# Understanding children’s behavioral health outcomes: A story of overcoming trauma and building strengths

**DOI:** 10.1101/2024.01.25.24301790

**Authors:** Jordan H. McAllister, Olga A. Vsevolozhskaya, Xiaoran Tong, Daniel P. Lakin, Scott K. Fairhurst, John S. Lyons

**Affiliations:** Center for Innovation in Population Health, University of Kentucky, Lexington, KY, USA; Pacific Clinics, Arcadia, CA, USA

**Keywords:** behavioral health, trauma, adverse childhood experiences, strengths, improvement, resilience

## Abstract

**Background:** Among children enrolled in behavioral health treatment, those with multiple trauma experiences (known as Adverse Childhood Experiences, or ACEs) typically see worse outcomes. In this study, we examine whether having or building strengths can help such children become more resilient and experience better outcomes.

**Objective:** We examined the relationship between children’s traumatic experiences, strengths, and clinical improvement, testing whether building strengths can help reduce the negative impact of ACEs on children’s response to treatment.

**Participants and Setting:** We used data from an evidence-based assessment to understand the clinical and functional needs and strengths of 5,423 children (ages 6-20) receiving treatment between 2019 and 2022 within a large community agency located in California.

**Methods:** To classify children by both level and rate of improvement, we relied on machine learning and principal components analysis. To determine the relationships between ACEs, strengths, and improvement, we used a variety of predictive models and descriptive analyses.

**Results:** After classifying children as being either “Faster”, “Slower”, or “Minimal” improvers, our analyses revealed that while higher total ACEs increases the likelihood of being a Slower improver, this effect can be mitigated by building strengths.

**Conclusions:** These results suggest that children with more ACEs are likely to require a longer duration of treatment before improvement is seen. They also suggest that promoting resilience— specifically focusing on building strengths—may lead to more efficient and effective care, particularly for children with significant trauma histories.

## Introduction

While there is widespread evidence that not enough youth who experience mental health challenges receive treatment (Merikangas et al., 2010; Wang et al., 2023), successful outcomes are not guaranteed even for those who do access specialty care. A wide range of studies, including meta-analyses and randomized clinical trials (Bear et al., 2020; Ng et al., 2023; Weisz et al., 2017), suggests that effect sizes from treatment range between moderate to low. Because of the typically lower impact of therapy for youth, there is a need for more research to investigate differences in improvement over time to see what changes tend to happen–and in what order and for whom (Ng et al., 2023).

Among those who receive care, children with multiple concurrent problems, who are often seen in community care, show the smallest effect sizes (Weisz et al., 2017). One study found that while just over half (54%) of youth treated in a Health Maintenance Organization (HMO) saw improvement, not quite half (44%) of those in a Community Mental Health Center were likely to improve (Warren et al., 2010). Possible reasons for variations across treatment settings include the increasing use of evidence-based practices (EBPs) in the HMO setting, differences in professional credentials, and social determinant complexities in the community setting.

In addition to these treatment disparities, a greater exposure to traumatic experiences for low-income children (Evans & Kim, 2013), many of whom are served in Community Mental Health Centers, further complicates the likelihood of successful treatment outcomes. Multiple studies have found that children who have Adverse Childhood Experiences (ACEs) are more likely as adults to suffer from issues related to substance use, mental health, physical health, and conditions like heart disease, cancer, lung disease, skeletal fractures, and liver disease (Evans & Kim, 2007; Felitti et al., 1998; Merikangas et al., 2009). Others have reported additional associations between ACEs and more frequent school absences (Lansford et al., 2002), worse quality of life (Afifi et al., 2007), dangerous sexual and criminal behavior (Gilbert et al., 2009), lower resilience (Collin-Vézina et al., 2011), more somatic complaints (Flaherty et al., 2013), and more severe risk behaviors, emotional and behavioral needs, and overall functioning (Kisiel et al., 2017).

Although ACEs are clearly associated with poor socioemotional, medical, and educational outcomes, the impact of these experiences is not uniform across people. This becomes apparent in the implementation of evidence-based trauma treatments, where the participants on whom the treatments were tested are notably less complex than those seen in clinics (Weiner et al., 2009; Weisz et al., 2013). In addition, the literature on resilience has identified certain protective factors that allow children to achieve better outcomes despite their traumatic experiences. One early study found that higher levels of peer, psychological, school/vocational, and moral/spiritual strengths all predict lower symptoms, risk behaviors, and functional impairment (Lyons et al., 2000). Other potential protective factors include resilient personality traits (Haskett et al., 2006), stable living situation (Bethell et al., 2014; DuMont et al., 2007), adaptive functioning skills (Schultz et al., 2009; Spinelli et al., 2023), family-level factors (Afifi & MacMillan, 2011; Moore & Ramirez, 2016), supportive relationships (Crouch et al., 2019), and a sense of purpose (Hamby et al., 2018).

Well-established practices designed specifically to treat symptoms of trauma—for example, Trauma-Focused Cognitive Behavioral Therapy (Cohen et al., 2016)—have inspired studies of the process of change that clients undergo on the road to improvement. For example, Ready and colleagues (2015) examined the degree to which clients receiving TF-CBT are able to decrease symptoms of overgeneralization by increasing accommodation, which the authors defined as “[t]he extent to which the person shows a balanced view of self, others, or the world.” The finding that clients who display accommodation are less likely to experience the negative impact of overgeneralization points toward developing a strength—as opposed to exclusively decreasing a symptom—as an important practice.

Given this well-known but complex relationship between traumatic experiences and behavioral health outcomes, recent work has emphasized the importance of creating a trauma-informed system of care (Ko et al. 2008; Lyons & Fernando, 2023; Oral et al., 2016). Such a system maintains records of traumatic experiences and traumatic stress, using that person-centered data to embed trauma-informed work in everyday practice at both the individual and system levels. Within the context of a large statewide agency committed to implementing such an approach, we investigated the effects of early trauma experiences on treatment process and outcomes to help deepen our understanding of likely trajectories of recovery for children. To do so, we used recent service utilization and Child and Adolescent Needs and Strengths (CANS) data on thousands of children from Pacific Clinics, California’s largest provider of behavioral and mental health services, and studied the relationship between traumatic experiences, strengths, and both clinical and functional improvement.

This study contributes to the existing literature by using a large, practice-based dataset to measure improvement over a comprehensive set of clinical and functional indicators, isolate the effects of ACEs and strengths, and directly test whether strengths are protective in the face of traumatic experiences. In this way, we attempt to answer two key questions. First, we seek to describe the relationship between ACEs and the trajectory of recovery in community-based care. Second, we seek to understand the role of strengths in recovery for children and youth with ACEs. Specifically, in this paper we built trajectories of recovery for children and youth in community-based care and study the relationship of ACEs with these different outcome trajectories. After determining the relationship between ACEs and outcomes, we identified what role, if any, strengths play in influencing these outcomes.

## Materials and Methods

### Data sources

Our primary source of information regarding trauma experiences, strengths, and clinical and functional outcomes was the Child and Adolescent Needs and Strengths (CANS). The CANS is an evidence-based, person-centered assessment that is the most widely used functional assessment for children and youth in the United States, and has a large body of published literature supporting its reliability and validity (Anderson et al., 2003; Kisiel et al., 2017; Lyons, 2022; Spinelli et al., 2023). Anyone administering the CANS is required to be certified with an interclass reliability passing score of at least 0.70. To guard against reliability decay, certification must be repeated annually.

The CANS assessment includes 58 indicators spanning the following domains: behavioral and emotional needs, caregiver needs and resources, cultural factors, life functioning, risk behaviors, transition to adulthood, strengths, and trauma. For each item in the CANS, children are given a rating between 0 and 3. For non-strength domains, an item rated ‘0’ means the child has no evidence of need and requires no action, ‘1’ means watchful waiting/prevention, ‘2’ indicates that the need is interfering with functioning and requires action, and a ‘3’ indicates a dangerous or disabling need that requires immediate/intensive action. For the strengths domain, a ‘0’ identifies one of the child’s centerpiece strengths that can be the focus of a plan, a ‘1’ indicates a strength that can be useful today, a ‘2’ indicates a strength that has been identified but needs to be developed before it is useful, and a ‘3’ indicates that no strength has been identified (Lyons & Fernando, 2021). For computationally intensive analyses, we dichotomized each CANS item as being either actionable (ratings of ‘2’ or ‘3’) or non-actionable (ratings of ‘0’ or ‘1’).

In addition to assessment data, demographic, diagnostic, and service utilization data was collected as a routine part of clinical care across participating clinical programs. The service data includes variables like type of service, type of program, date of service, and discharge reason.

### Working sample

Included in the sample were children between the ages of 6 and 20 who received services from Pacific Clinics between 2019 and 2022. This agency serves more than 50,000 clients annually, and provides services in nearly 20 counties across California. More than 96% of clients’ services are funded by MediCAL (California’s Medicaid program), and service provision catchment areas encompass both rural and urban regions. Service provision includes a range of services, such as prevention and early intervention, crisis support and stabilization, outpatient mental health services, foster care and adoption, and others. Youth included in the primary sample ranged in symptom acuity, diagnoses, and presenting problems.

The only criteria for inclusion in the sample were that each child must have both CANS and service data and at least two CANS assessments during that period. However, any child who had either all ‘0’s or ‘3’s across the strengths items for all CANS assessments was excluded from the sample when running analyses on strengths, as this suggests their strengths were not being assessed accurately.

We addressed missing data on individual items in the CANS by replacing any missing strengths values with ‘3’s (indicating no evidence of strength) and any other missing CANS values with ‘0’s (indicating no evidence of need). Along with the service data, we addressed the problem of repeat enrollment IDs (which are supposed to be unique for each assessment/service and each child) by creating a new ID that combines the enrollment ID with the date of assessment/service. Since each new ID may correspond to multiple CANS recorded in the same day for a child, we collapsed multiple rankings by taking the minimum for an item of strength, or the maximum for an item of need—capturing the highest strengths or needs a child had—and then removed duplicates.

This resulted in a sample of 5,423 children, each of whom had multiple CANS assessments and services. The average age of the children in the sample (at the time of entry into the system) is around 12, while there is an equal split between male and female children. The majority of children in the sample are Hispanic (58%), with white (13%), black (8%), Asian (6%), multiethnic (6%), and other (9%) ancestry following in that order.

### Measures

Across the various analyses, our main variables from the CANS included ten trauma items (medical trauma, natural disaster, criminal act, emotional abuse, physical abuse, community violence, sexual abuse, family violence, neglect, and disrupt caregiving) and nine strengths items (family, talents/interests, resiliency, natural supports, interpersonal, educational, community life, spiritual/religious, and cultural identity), from which we created measures of total ACEs (a count of all types of trauma experiences to which a child was ever exposed) and maximum strengths (the overall count of strength types a child ever built). We also relied on measures of length of time (number of 6-month intervals between first and last service date), demographic variables (race, sex, and age), counties (12 total across California), diagnoses (a binary indicator for each condition if ever diagnosed), and service intensity (average number of services received per month).

For the purposes of this study, we created a measure of disengagement. This measure accounts for the possibility that children are either not showing up or are not fully engaging in their services, whether that be because youth are checked out or because their caregivers fail to provide transportation to scheduled sessions (Hawley & Weisz, 2005). Succinctly, our disengagement measure represents the proportion of total services for which each child is disengaged. The following services are all considered disengaged: those labeled “no show” or including a discharge reason like “disengaged/withdrawal without all goals achieved: client decision”, “drop-out/lack of participation”, “violated program rules”, or “no pr[o]gr[e]ss”. We used disengagement both as a control in our main models and as an outcome variable when testing the protective effects of strengths against ACEs, given that our main outcome involves multiple improvement classes and so cannot easily be depicted along a single dimension.

In addition, we created a measure of improvement by combining multiple CANS items along with length of time services were received. This particular measure is one of the innovations of this piece and so will be described in more detail in the following sections.

### Analyses

It is necessary to carefully define “improvement”, because the most important aspect of outcomes for children enrolled in behavioral health services is their relative amount and speed of improvement. This is difficult to do, especially when considering the multifaceted nature of improvement and resilience (Ager, 2013; Hamby et al., 2018; Herrman et al., 2011; National Scientific Council, 2015). And while there is a large literature showing that psychotherapy generally leads to a moderate amount of improvement among children (Bear et al., 2020; Ng et al., 2020; Weisz et al., 2017), we are not aware of any existing work that considers the level and speed of improvement simultaneously.

The full CANS assessment consists of 58 items that cover trauma, strengths, and other factors relevant for improvement. Thus, each child is represented by 58 distinct time series, each tracking changes in individual item ratings. Within an item, we defined improvement as a change between *ever* actionable item and the *latest* actionable score. For example, if a child was ever actionable on an item (had a rating of 1) but had a non-actionable 0 rating on the most recent assessment, they have a positive within-item improvement rating of 1 – 0 = 1. If a child was ever actionable on an item and was still actionable on that item during the most recent assessment, they have a 1 – 1 = 0 improvement rating. If a child was never actionable on an item but became actionable during the most recent assessment, they failed to improve and have a negative 0 – 1 = – 1 rating. Hence, within items we defined “improvement” as a {-1, 0, 1} rating, with positive values indicating a positive change.

However, the overall level of improvement across all 58 items is not as easy to define. A seemingly simple solution for defining an overall improvement as a sum across 58 {-1, 0, 1} within-item improvement scores would be inappropriate because item ratings are correlated and the change of one item can anticipate the change of other items, since a person’s improvement (or lack thereof) is usually multifaceted. Additionally, observed within-item changes are expected to be highly correlated with the length of time services are received, which is also not going to be represented by a simple sum. Consequently, we quantified improvement across 59 dimensions (58 CANS items and the length of time services were received) by utilizing machine learning clustering techniques to put the data into distinct trajectories or classes.

Specifically, we assigned 5,423 rows (number of children) by 59 columns of data into a set number of classes. A priori, the classes are unknown and so are learned from the data. There are a variety of ways to do this using unsupervised machine learning. One such technique is a model-based clustering algorithm called Finite Gaussian Mixture, which identifies different classes in the data based on maximum likelihood estimation (Hartigan, 1985; McLachlan et al., 2019; Oberski, 2016). We used Finite Gaussian Mixture (FGM) because (1) it allows more flexible shape of the clusters in the 59-dimensional space, and (2) the distribution of values in each column is roughly bell shaped. The data largely follows a normal distribution because for each of the 58 CANS items, the majority of children have a value of 0 (i.e., neither improved nor worsened by the end of their treatment) while only a few have a value of 1 or –1. The one remaining column representing length of time is mostly concentrated between 6-12 months and so has a bell-shaped distribution as well. When fitting the FGM model, the only required hyper-parameter is the number of groups by which the data should be divided; we focused on the three-class grouping given that it produces the clearest separation while also being the most parsimonious solution.

We evaluated the validity of the three-class solution by examining how similar are changes across 59 variables (58 CANS items and the length of time) among children within the same class. Visual exploration is an essential component of data analyses, as it allows for the most intuitive evaluation of model performance. To visualize similarities across 59 variables, we turned to principal component analysis. Principal components analysis (PCA) is a dimension reduction technique used to visualize multivariate data. The four main goals of PCA include (1) extracting as much information from the data as possible; (2) reducing the size of the data by keeping only the most important information; (3) simplifying the description of the data; and (4) analyzing the structure of the data (Saporta & Keita 2009).

In our analysis, we visualized children’s CANS data by reducing its 59 dimensions to only two (the first and second principal components). Each principal component (PC) is a linear combination of the original variables in the dataset, where the first PC explains the most variation in the data. The second PC explains the most variation residual to the first PC (i.e., remaining variation in the data after removal of information explained by the first PC), to ensure that it is not capturing redundant information (Abdi & Williams, 2010). By plotting the data across the first two PCs, we showed how the data is distributed across those 58 different CANS items (plus a variable for length of time) and how similar children within each class are to one another.

To determine the impact of ACEs on improvement, we used both LASSO and logistic regressions. We fit a multinomial LASSO model on the entire data, using the three assigned class labels as the outcome variable, total ACEs as the explanatory variable, and the following variables as controls: age, sex, race, county, diagnoses, service intensity, disengagement proportion, and all CANS items other than ACEs. We also subsetted to only Faster and Slower improvers before running a logistic model to predict whether children will see slower (rather than faster) improvement based solely on their total ACEs (and then reran it but including controls).

Additionally, we conducted analyses investigating how strengths vary over time among the different classes, as well as testing whether strengths attenuate the correlation between ACEs and worse outcomes–in this case, disengagement. To look at strengths over time, we split the initial and final CANS assessments into approximately equal-sized groups of either “high” or “low” strengths and then compared the proportions of each improvement class that have high initial versus final strengths. And to investigate how strengths interact with ACEs to affect disengagement, we plotted the data by total ACEs (x axis) and disengagement (y axis) but divided into groups by total strengths.

## Results

### Defining improvement

Figure 1 shows the data points of 5,423 children projected onto the first two PCs (see Appendix 1 for the PC loadings), colored by their FGM-assigned class (see the scatter plot and two neighboring density plots), as well as the relative size of each class (see the bar chart) and total ACEs by class (see the boxplot).

**Fig. 1.**
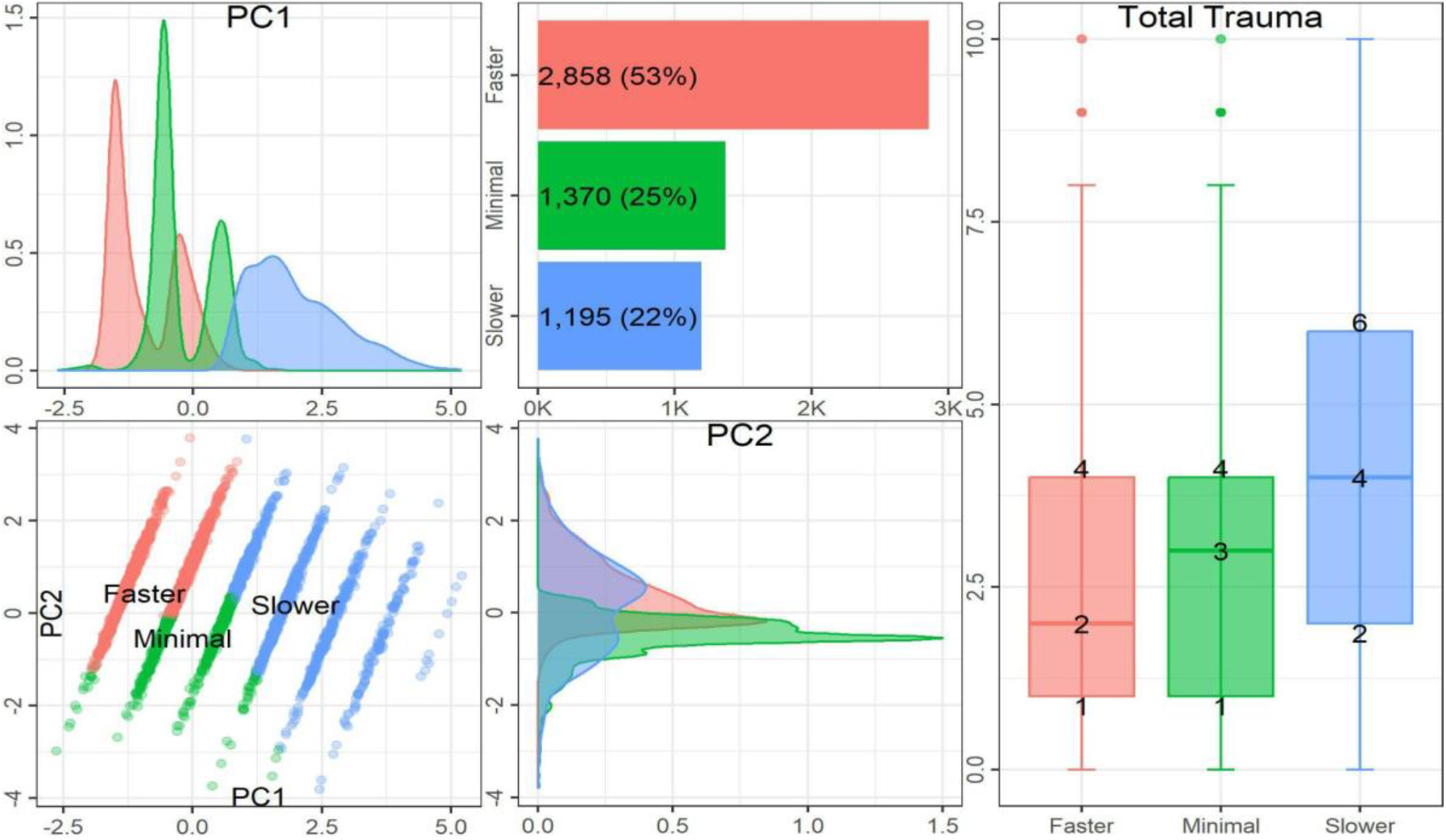
This figure uses PCA to portray the results of the FGM. The bottom-left scatter plot shows the data (labeled by class) plotted by PC 1 and PC 2, where the density plots for all three classes are above (PC 1) and to the right (PC 2). The bar chart at the top displays the relative size of each class, and the boxplot on the right captures the interquartile range of total ACEs for each class.

Focusing on the scatter plot, PC 1 looks unusual, as the data is strictly divided into lines along the x axis. This is because the first PC is composed mainly of the length of time services were received (which has a loading of 0.94), meaning this particular measure explains the most variation in the data. This indicates that length of time in treatment is strongly related to a person’s overall improvement. Since our length of time variable is coded in 6-month intervals, the visualization produces a single line for each of the 6 months.

The second PC (the y axis) is a combination of many different CANS items—with the highest loadings (or correlations between the components and variables) being for family functioning (loading = 0.27), interpersonal strength (loading = 0.25), resiliency (loading = 0.24), and social functioning (loading = 0.24)—that transition from actionable to non-actionable (and so “improve”) moving up along the axis. All CANS items’ loadings for PC 2 are assigned a positive value (given that improved items are coded as ‘1’) except for some of the trauma items (sexual abuse, physical abuse, natural disaster, family violence, community violence, and criminal acts), which are assigned a loading of 0. So whereas PC 1 captures length of time in services, PC 2 captures children’s improvement while in services. After length of time, this particular combination of variables explains the second-most variation in the data.

Figure 1 also shows the grouping of data as classes derived from FGM. The three classes identified are distinct and interpretable when graphed along the first two PCs. The largest class is the pink group (labeled “Faster”), which contains children who improve by a lot over a short period of time. The second-largest class is the green group (labeled “Minimal”), which includes children who see little improvement over a short period of time. The smallest class is the blue group (labeled “Slower”), which comprises children who see a range of improvement levels (but typically improve more than the Minimal class) over a longer duration. In this way, each class accounts for both the children’s relative level of improvement and their speed of improvement.

### Determining impact of trauma

In our LASSO model (see Appendix 2 for the full results), total trauma (ACEs) is positively associated with Slower improvement (r = 0.2) and negatively associated with Faster improvement (r = –0.07). In other words, a child with a greater number of ACEs is more likely to be a Slower improver and less likely to be a Faster improver. The associations between ACEs and Faster versus Slower improvement is portrayed in Figure 2, which depicts the relationship between children’s total number of ACEs and their likelihood of falling into the Faster, Slower, or Minimal improvement classes.

**Fig. 2.**
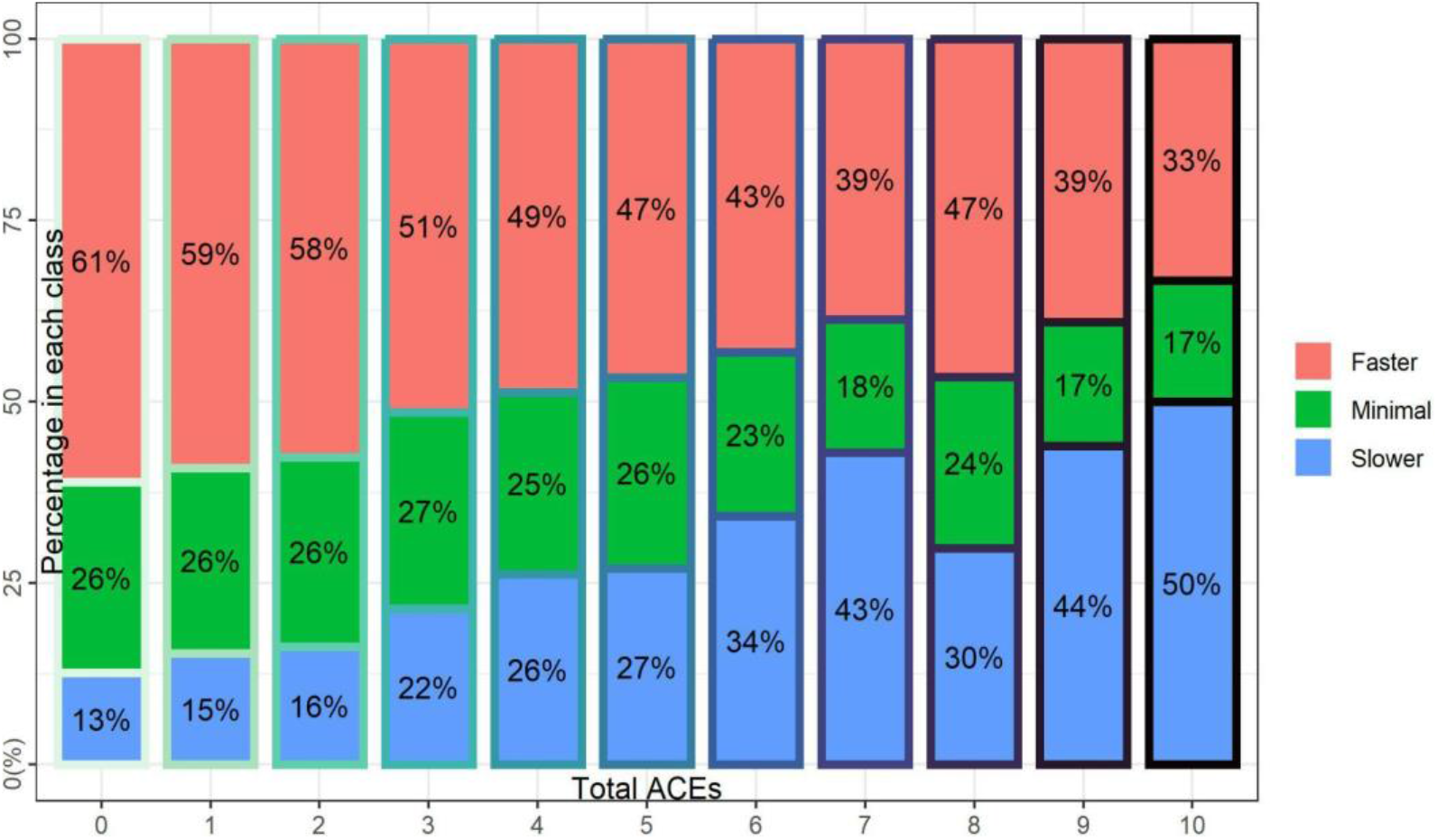
This stacked bar plot shows the relative percentages by class (y axis) for each level of total ACEs (x axis).

Figure 2 portrays the overall percentage of children within the three classes at each level of ACEs. The graph makes it evident that there is a strong linear relationship between total ACEs and the likelihood of Faster versus Slower improvement. With each additional ACE, the likelihood of being in the Faster class goes down while the likelihood of being in the Slower class goes up. There is not a plateau effect after a certain number of ACEs, and instead each ACE matters. This reflects studies that have found a dose-response effect, in which outcomes worsen incrementally with every new ACE (Bethell et al., 2014; Felitti et al., 1998; Flaherty et al., 2013; Goldenson et al., 2020).

Figure 3 depicts a logistic model (see Appendix 3 for the full results) capturing the relationship between total ACEs and the likelihood of being a Slower rather than a Faster improver (r = 0.2***).

**Fig. 3.**
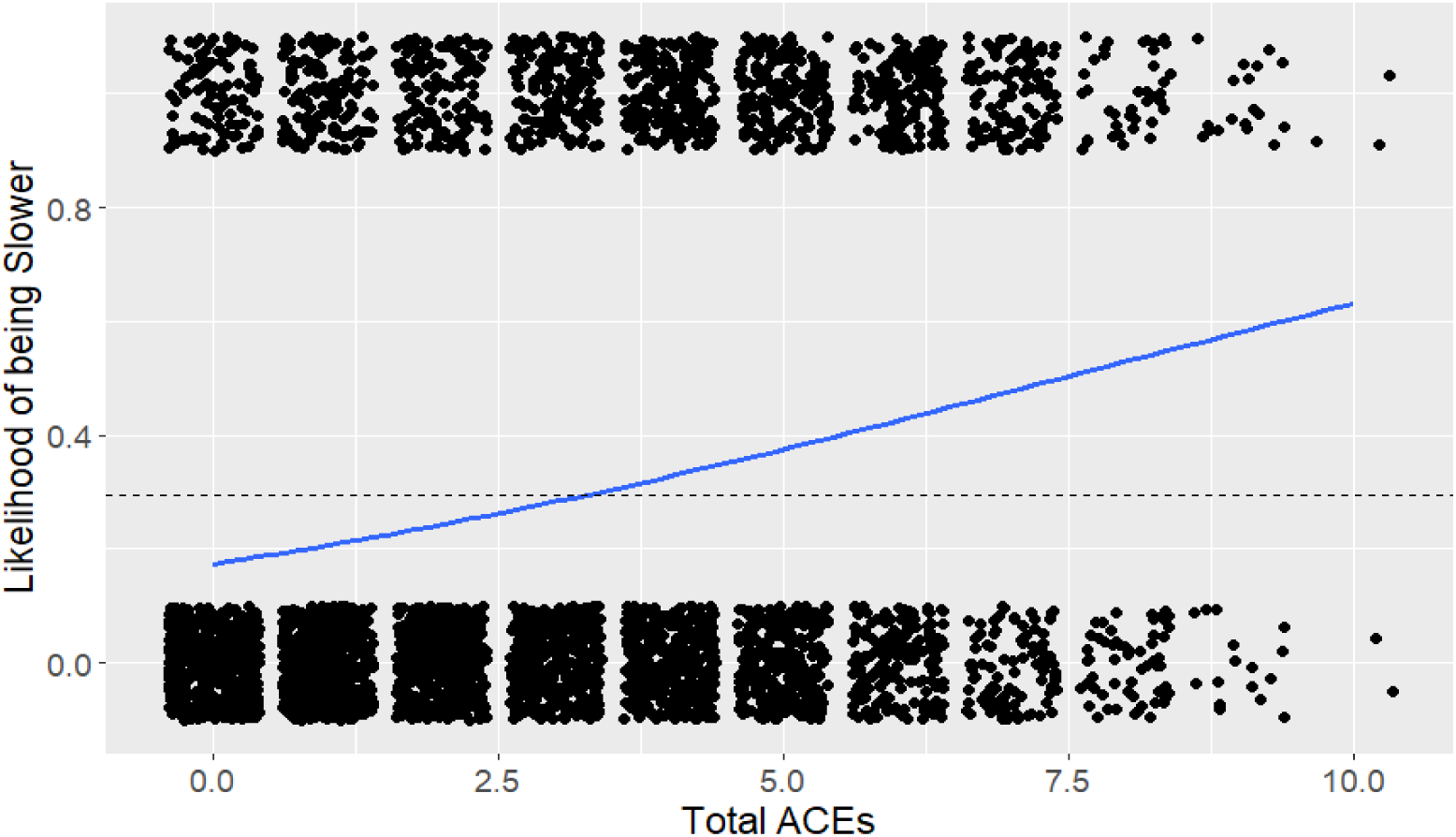
A logistic regression line captures the effect of total ACEs (x axis) on the likelihood of being a Slower versus Faster improver (y axis). The dotted line represents the proportion of Slower improvers in the entire sample, and all Slower and Faster improvers’ total ACEs are plotted at either the top (Slower) or the bottom (Faster) of the graph.

In this graph, the regression line crosses the dotted line (representing the proportion of Slower improvers in the overall sample) after approximately 3 ACEs. So once they have more than 3 ACEs, children become more likely to be Slower rather than Faster improvers. This result can be deduced even just by observing the data points plotted for both the Slower (see top of graph) and Faster (see bottom of graph) improvers. The highest density of Slower improvers occurs between 4-6 ACEs, whereas the highest density of Faster improvers occurs between 0-3 ACEs.

Rerunning the same logistic regression but including controls for other factors that can influence improvement produces Figure 4 (see Appendix 3 for the full results). Like before, r = 0.2***, but notice that the y-intercept is lower.

**Fig. 4.**
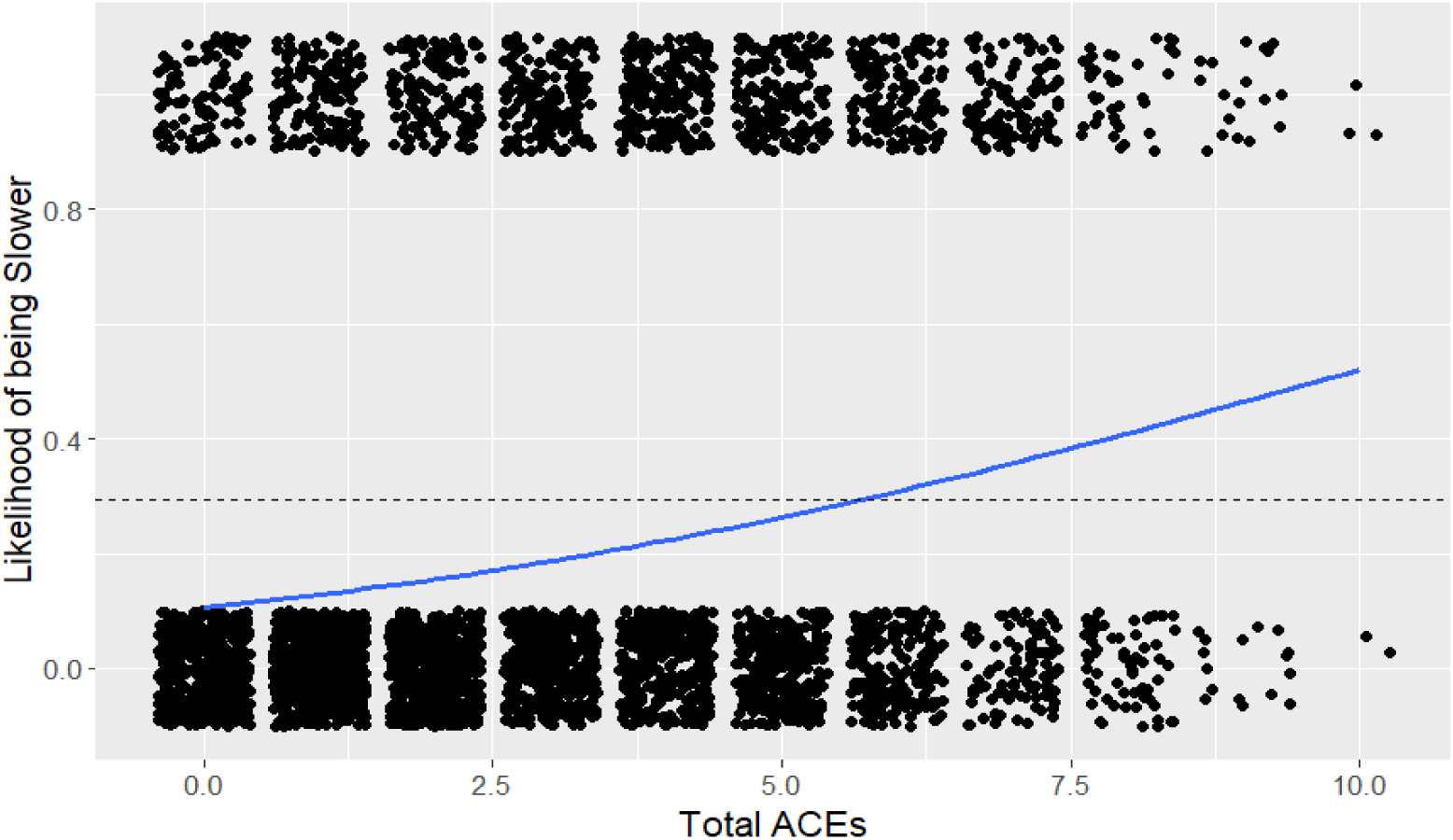
A logistic regression line captures the effect of total ACEs (x axis) on the likelihood of being a Slower versus Faster improver (y axis), controlling for age, sex, race, county, diagnoses, and services. The dotted line represents the proportion of Slower improvers in the entire sample, and all Slower and Faster improvers’ total ACEs are plotted at either the top (Slower) or the bottom (Faster) of the graph.

Figure 4 differs from Figure 3 only in that the regression line now crosses the sample proportion of Slower improvers after approximately 5 total ACEs. So as we might expect, the inclusion of other relevant variables (such as diagnoses and services) attenuates the effect of ACEs on improvement. Still, once they have more than 5 ACEs, children become likely to be Slower rather than Faster improvers—even when controlling for these other factors. Therefore, the threshold for high levels of trauma appears to fall somewhere between 3 to 5 total ACEs. This reflects various studies using 4 ACEs as the cutoff for highly traumatized children (Crouch et al., 2019; Felitti et al., 1998; Goldenson et al., 2020; Lyons & Fernando, 2023).

### Determining impact of strengths

For children with high initial strengths (a sum strengths value less than or equal to 9, meaning they had mostly ‘0’s or ‘1’s for all nine strength items on their initial CANS assessment), 52% are Faster improvers, 19% are Slower improvers, and 29% are Minimal improvers. For the high final strengths group (a sum strengths value greater than or equal to 9 on their final CANS assessment), 56% are Faster improvers, 23% are Slower improvers, and 21% are Minimal improvers (see Appendix 4 for a table with all the percentages). While the proportion of Faster and Slower improvers increases by 4% each between the high initial and final strengths groups, the proportion of Minimal improvers decreases by 8% between the high initial and final strengths groups. Thus, Faster and Slower improvers are building strengths over time, whereas Minimal improvers are not.

Figure 5 illustrates the relationship between trauma, strengths, and disengagement.

**Fig. 5.**
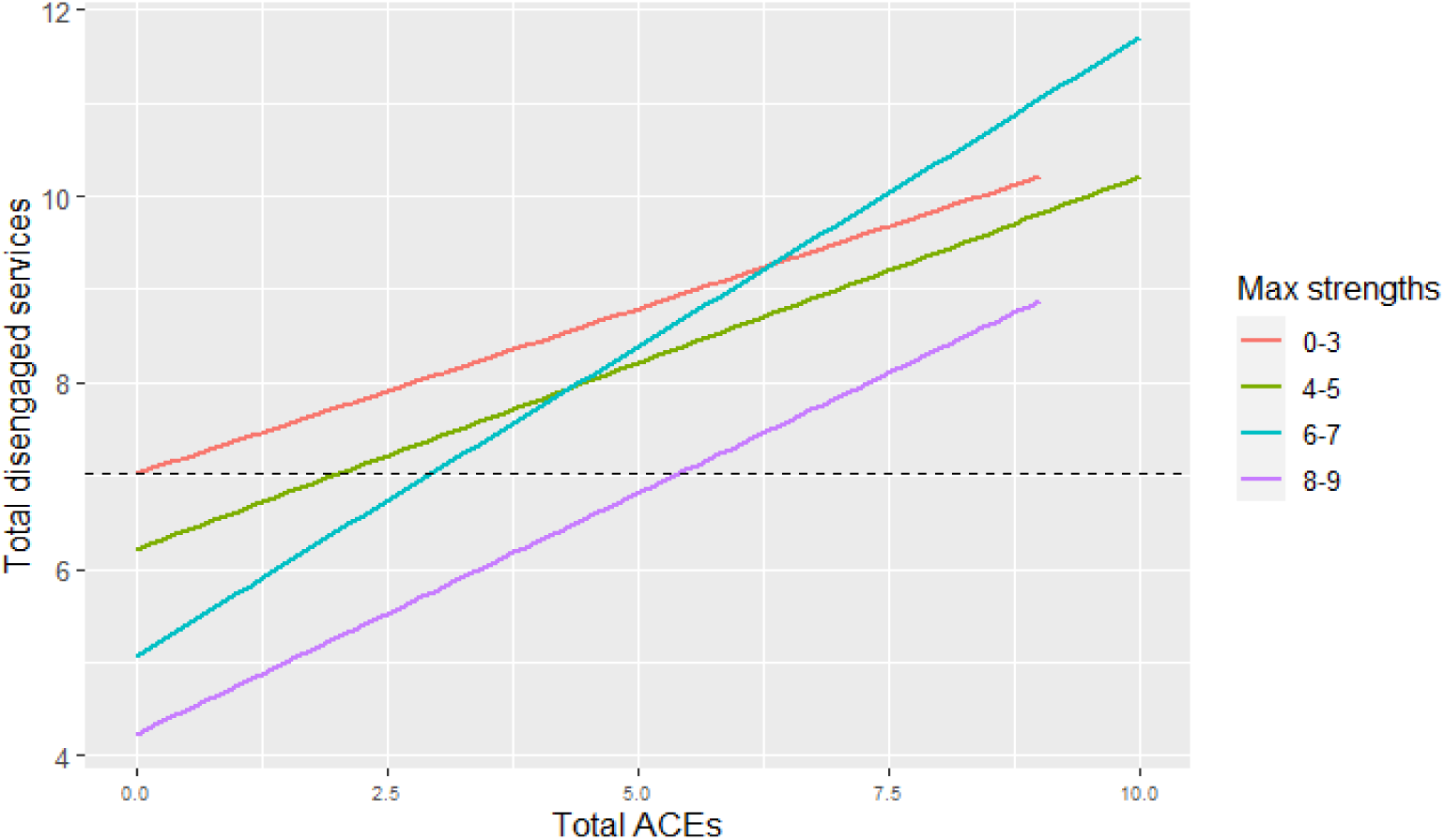
The four colored lines capture the relationship between total ACEs (x axis) and total disengaged services (y axis) for children with 0-3, 4-5, 6-7, and 8-9 maximum total strengths. The dotted line represents the average number of total disengaged services in the entire sample.

Here, the data is divided into four groupings by maximum total strengths, represented by the different color lines. Children with only 0-3 strengths (red line) start out at the average total number of disengaged services (represented by the dotted line) even if they have no ACEs. Then with each additional ACE, their level of disengagement only increases from there. By contrast, children with 8-9 strengths (purple line) are predicted to have a lower-than-average number of disengaged services at 0 ACEs, and it is only after approximately 5 ACEs that they reach the same average level of disengagement that the lowest strengths group started out with. This same mitigating effect carries over across all the strengths groups, where higher strengths groups require more ACEs to see the same worse outcomes as lower strengths groups.

## Discussion

This study applied statistical innovations to clinical data to provide insight into how youth, particularly those with a history of complex trauma, improve during mental health treatment. The literature has shown that a history of childhood trauma experiences predicts worse clinical and functional outcomes while strengths predict better outcomes; however, it is less clear whether strengths are protective against the negative effects of trauma experiences in terms of treatment outcomes. The results of the present study shed light on this interplay of trauma and strengths in predicting and, perhaps, influencing outcomes for children receiving community-based behavioral health interventions.

We gained key insights by using a large, person-centered dataset that includes clinical and functional measures for children enrolled in services provided by California’s largest community behavioral health agency. We employed FGM to cluster the data into distinct classes, as well as PCA to visualize the data along two dimensions. PCA displayed the data along PC 1 (mainly composed of length of time in services) and PC 2 (a combination of multiple CANS items, all of which are improving). This allowed us to see how the data is divided by both length of time and improvement, and thus we were able to define the three classes identified by FGM as being either “Faster”, “Slower”, or “Minimal” improvers.

When comparing levels of improvement across children enrolled in behavioral health services, the importance of ACEs and strengths is evident. ACEs are positively associated with Slower improvement and negatively associated with Faster improvement. This indicates that children with a higher number of ACEs (specifically, greater than approximately 4 ACEs) are more likely to require a longer duration of treatment in order to see improvement. The higher the traumatic experience “loading”, the longer it will take to achieve good clinical and functional outcomes. Importantly, children with higher loadings still have positive outcomes—it just took longer to achieve. Service providers should take this into account when conducting utilization management, as certain children (i.e., those with more trauma experiences) can be expected to require a longer duration of treatment before improvement is seen.

Regarding clinical opportunities, we have shown that strengths increase over time for children who improve–whether that improvement be fast or slow–but not for those youth who do not. In examining how ACEs and strengths together affect disengagement with treatment, we found that children with many strengths are predicted to have above-average levels of engagement even with relatively high levels of ACEs. Speaking directly to the protective role of strengths, this finding suggests that strengths may, in fact, reduce the negative impact of ACEs on children’s participation in and their outcomes within community-based behavioral healthcare. This represents an opportunity to add to both the evidence base and to a clinical rationale for strength-building as a legitimate treatment activity.

While this investigation supports the role of resilience in both successful treatment engagement and outcomes, future avenues for research should continue to explore the potential predictive nature of specific strengths, needs, and clinical factors as they relate to the likelihood of continued involvement in mental health services. Given the paucity of consistently effective and accessible treatment for children living in poverty, practice-based investigations that allow for more accurate identification of both treatment response and rate of response could lead to more consistent and effective treatment for high-risk youth. It appears likely that a focus on strength identification and building early in treatment is a promising strategy in this context.

## Limitations

As with any assessment, the CANS may not always fully capture all relevant variables. Although the children are evaluated by trained CANS assessors, some information may be withheld, overlooked, or entered incorrectly. There may even be certain types of trauma experiences and strengths that are not included in the CANS, which would therefore fail to provide that information. And when cleaning this dataset as well as the service data provided by Pacific Clinics, issues of missing data, repeated observations, and inconsistencies were addressed systematically, but certain information could have been lost or limited by the cleaning process.

Given the nature of our data, it can be argued that these findings may not be generalizable to other time periods, agencies, regions, or clients whose treatment is privately funded. As our data spans the period between 2019 and 2022, there may be some concern that these findings were affected by the COVID-19 pandemic and so may not carry over to subsequent years. And as Pacific Clinics is based in California, these findings may not be applicable to other agencies and regions. Treatment of clients served by Community Mental Health Centers is largely funded through Medicaid and is therefore more likely to represent clients with more complex trauma histories than, for example, clients treated through HMOs. Confirmation of these relationships, of course, requires other researchers to conduct similar analyses using data from other providers, funding sources, and regions of the United States, to see whether they find similar effects.

Additionally, the use of machine learning is data-specific in that the specific three-class solution and two principal components are based exclusively on the data provided by Pacific Clinics. It is unclear whether similar classes and principal components would be found if using another dataset. It could even be the case that a three-class solution does not provide the clearest separation when using another provider’s data, which may require more classes to best describe trajectories of recovery.

## Conclusion

The results of this study strongly recommend that building strengths (and thus promoting resilience) leads to better and more efficient outcomes of community-based behavioral health treatment, especially for children with multiple ACEs. This is a notable finding, given that most treatment protocols and utilization management strategies continue to focus on the elimination of symptoms as the primary activity that leads to improvement. While it is important to consider all aspects of recovery, particularly as it applies to effective clinical interventions, the analyses contained in this paper consistently point to the importance of building strengths. Indeed, an effective trauma-informed system of care must not only record and treat trauma, but also identify and work on areas in which strengths can be developed, thus increasing the resilience and consequent improvement of patients with high levels of trauma.

## Data availability

Given the sensitive nature of the data, it is not able to be released upon publication of this paper.

## Acknowledgements

Many thanks to various team members at the Center for Innovation in Population Health and Pacific Clinics for their helpful feedback. We also appreciate the suggestions offered by participants in the 2023 TCOM conference held in Lexington, Kentucky.

## Appendix 1. Principal component loadings

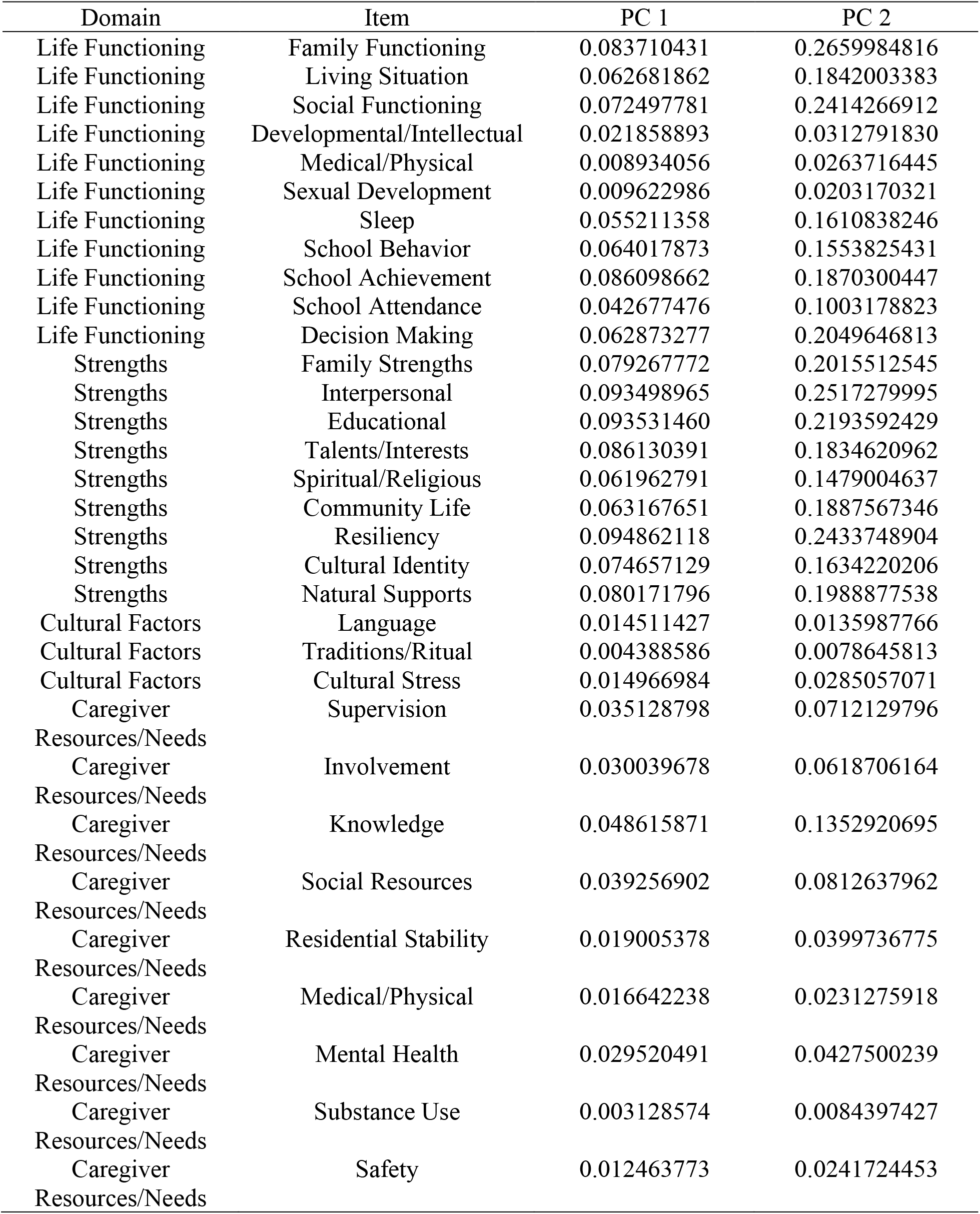
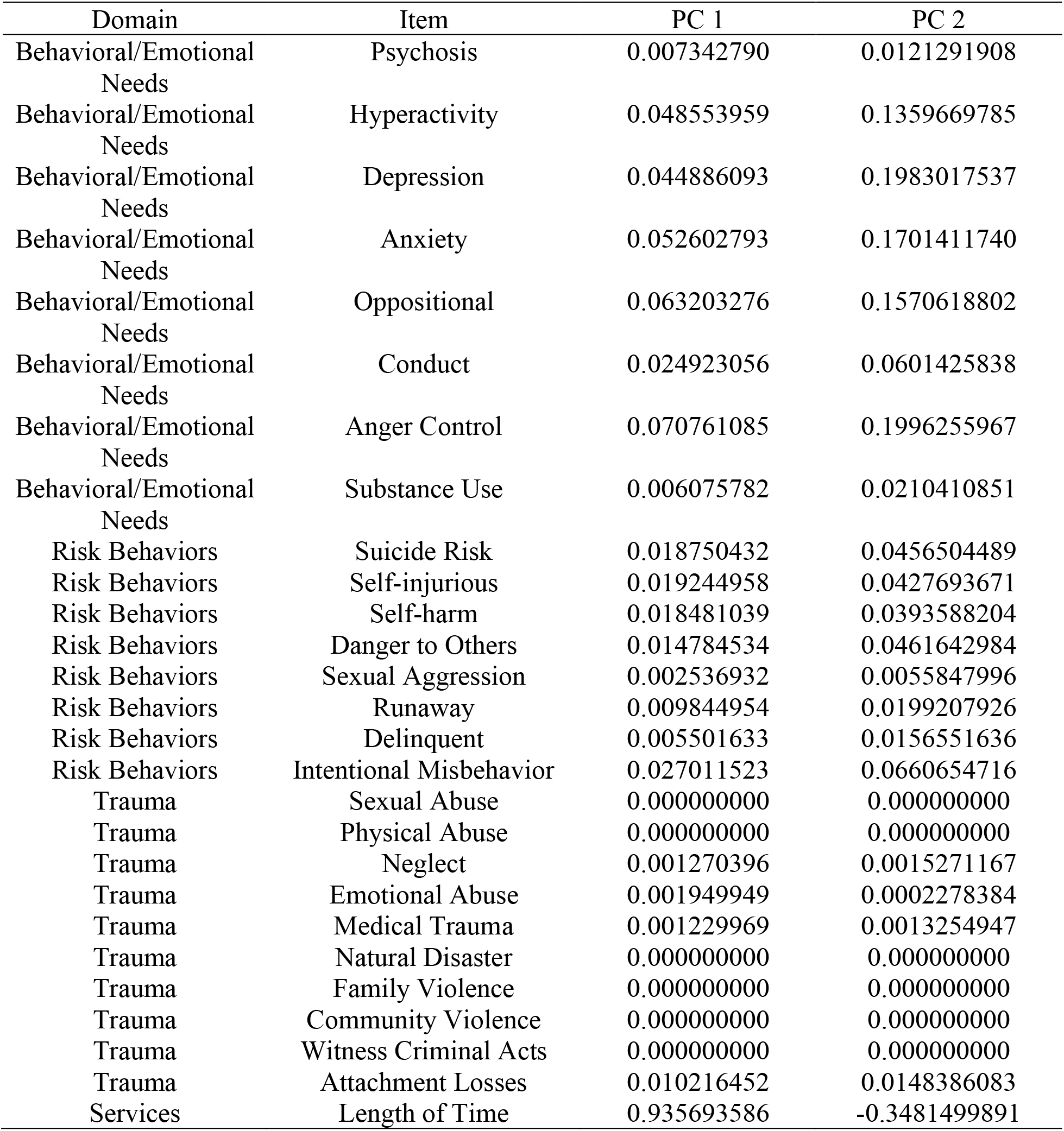

## Appendix 2. LASSO model results

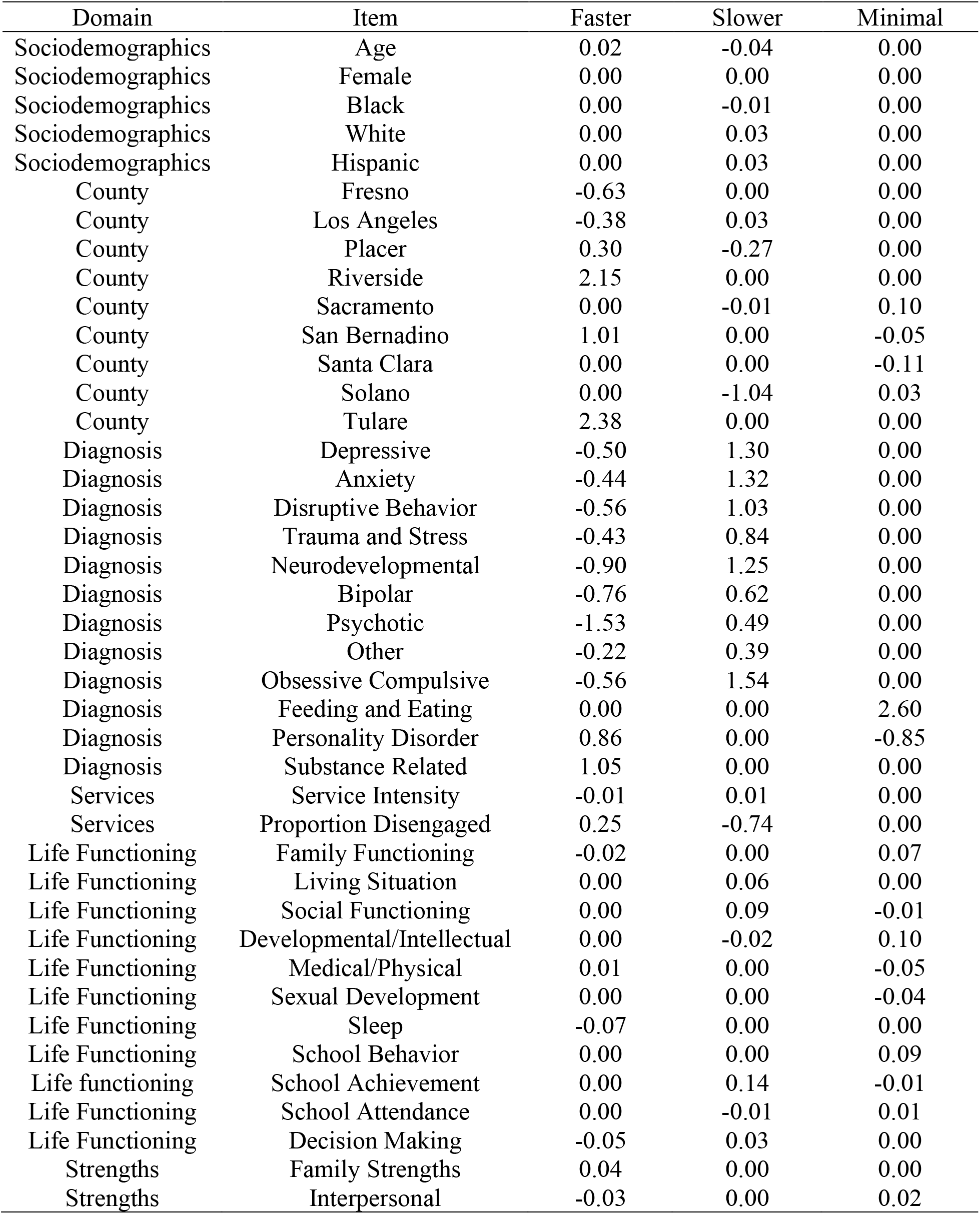
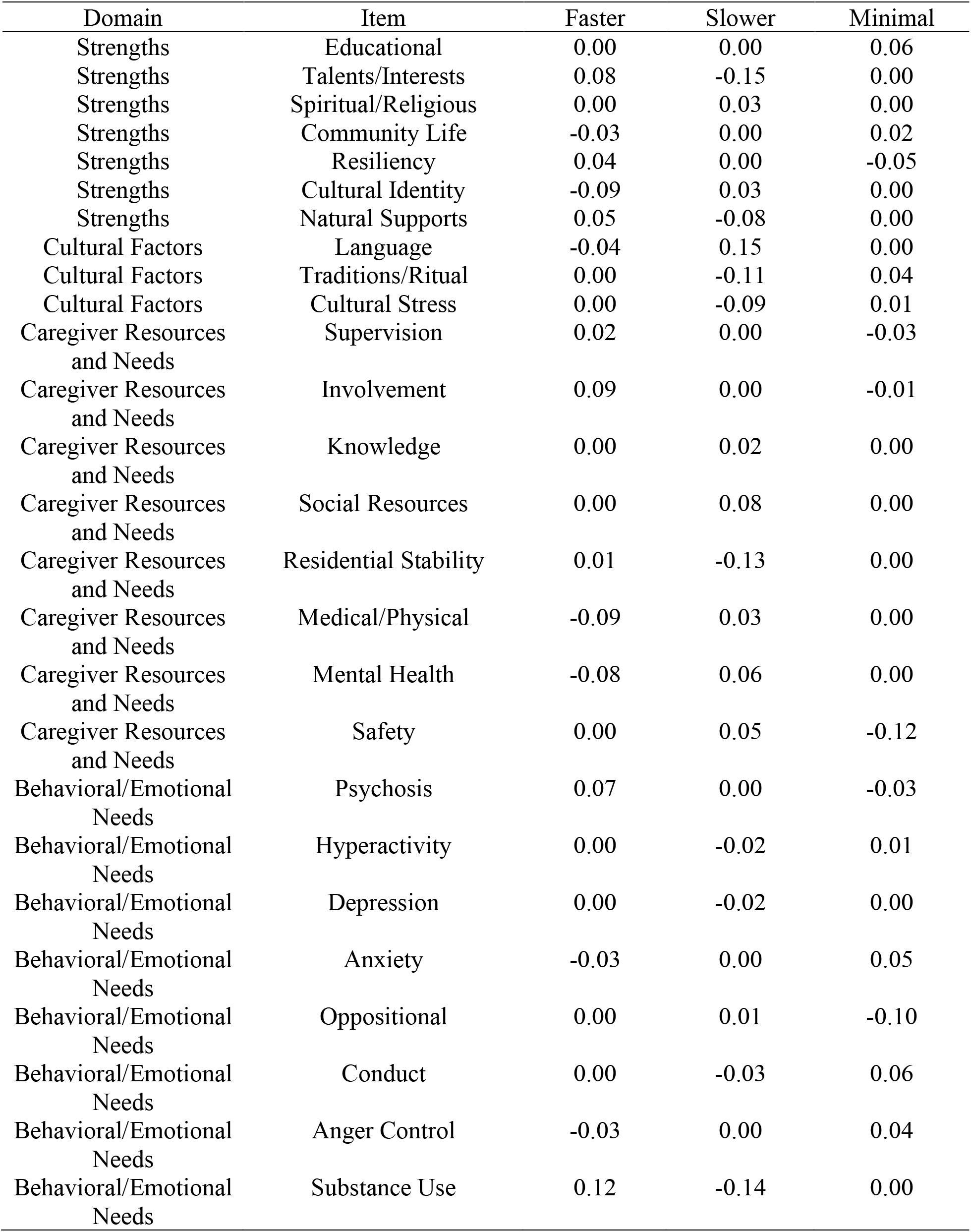
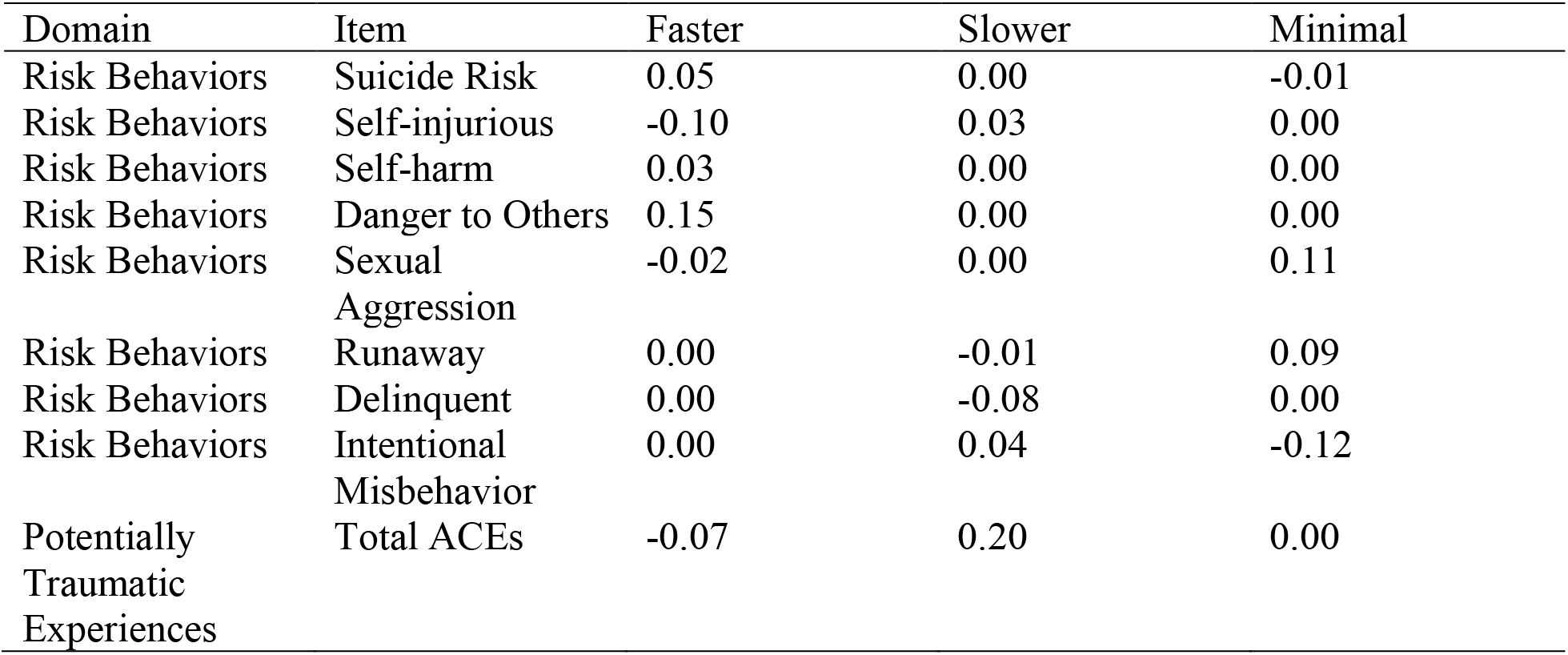

## Appendix 3. Logistic regression results

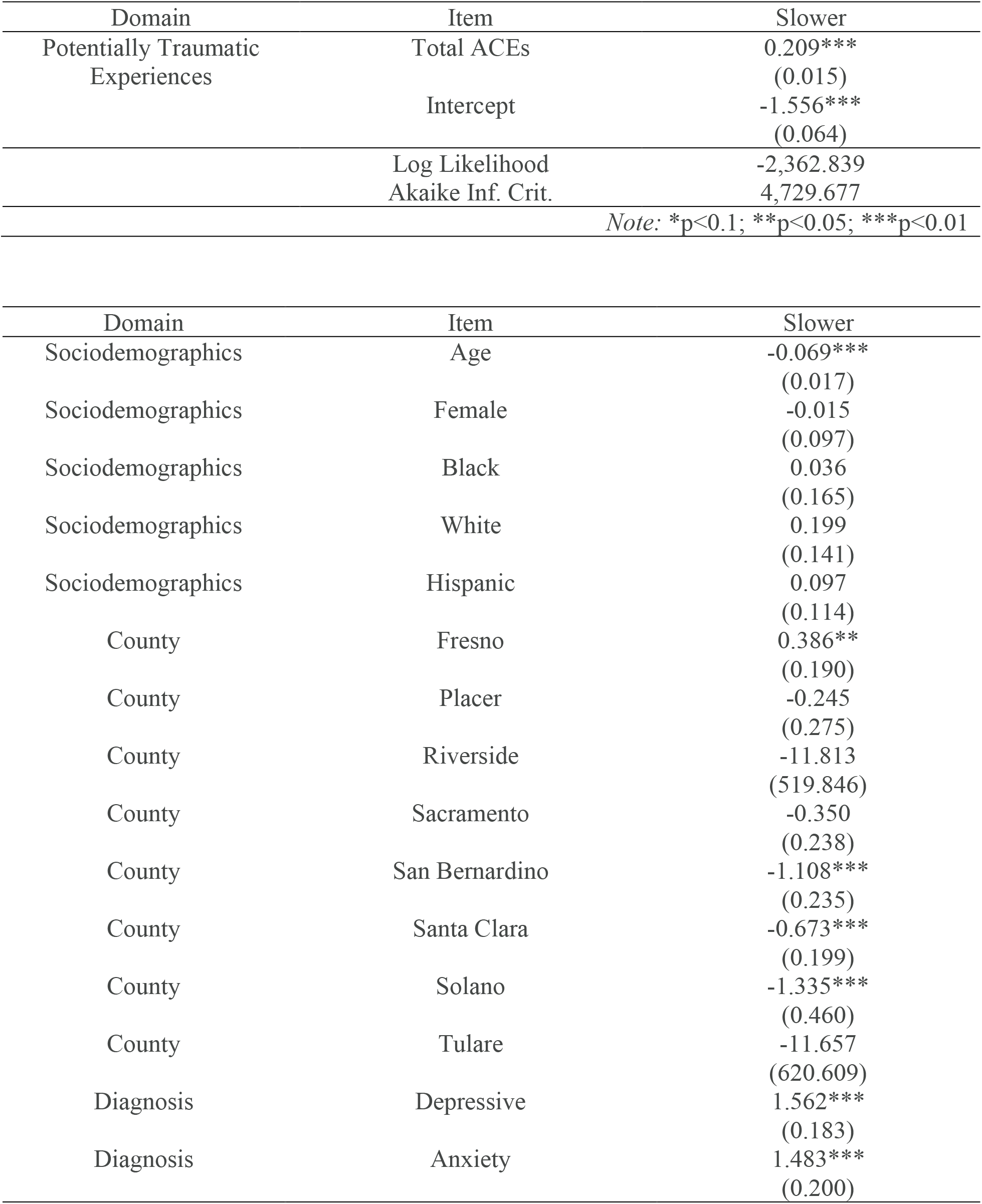
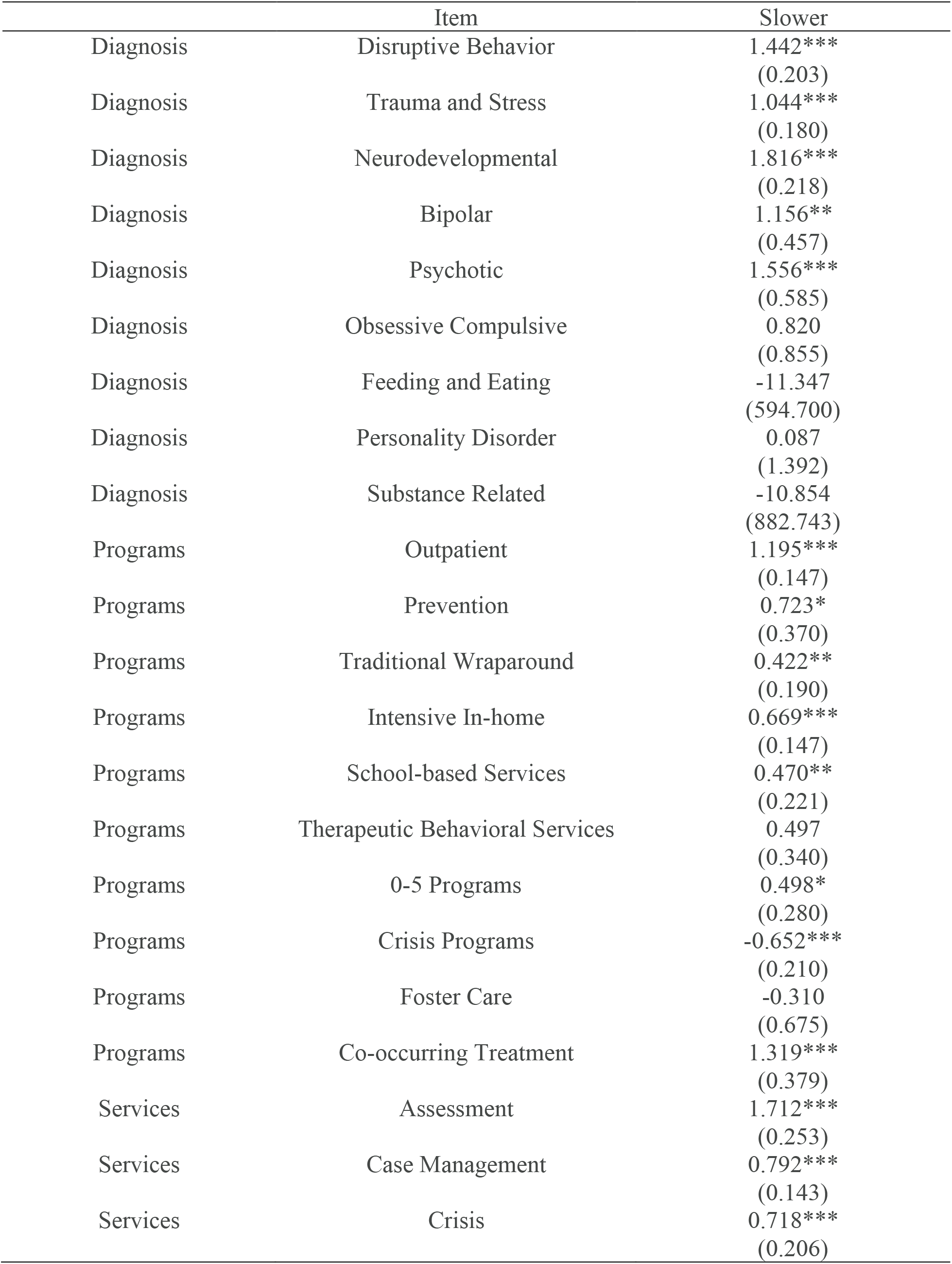
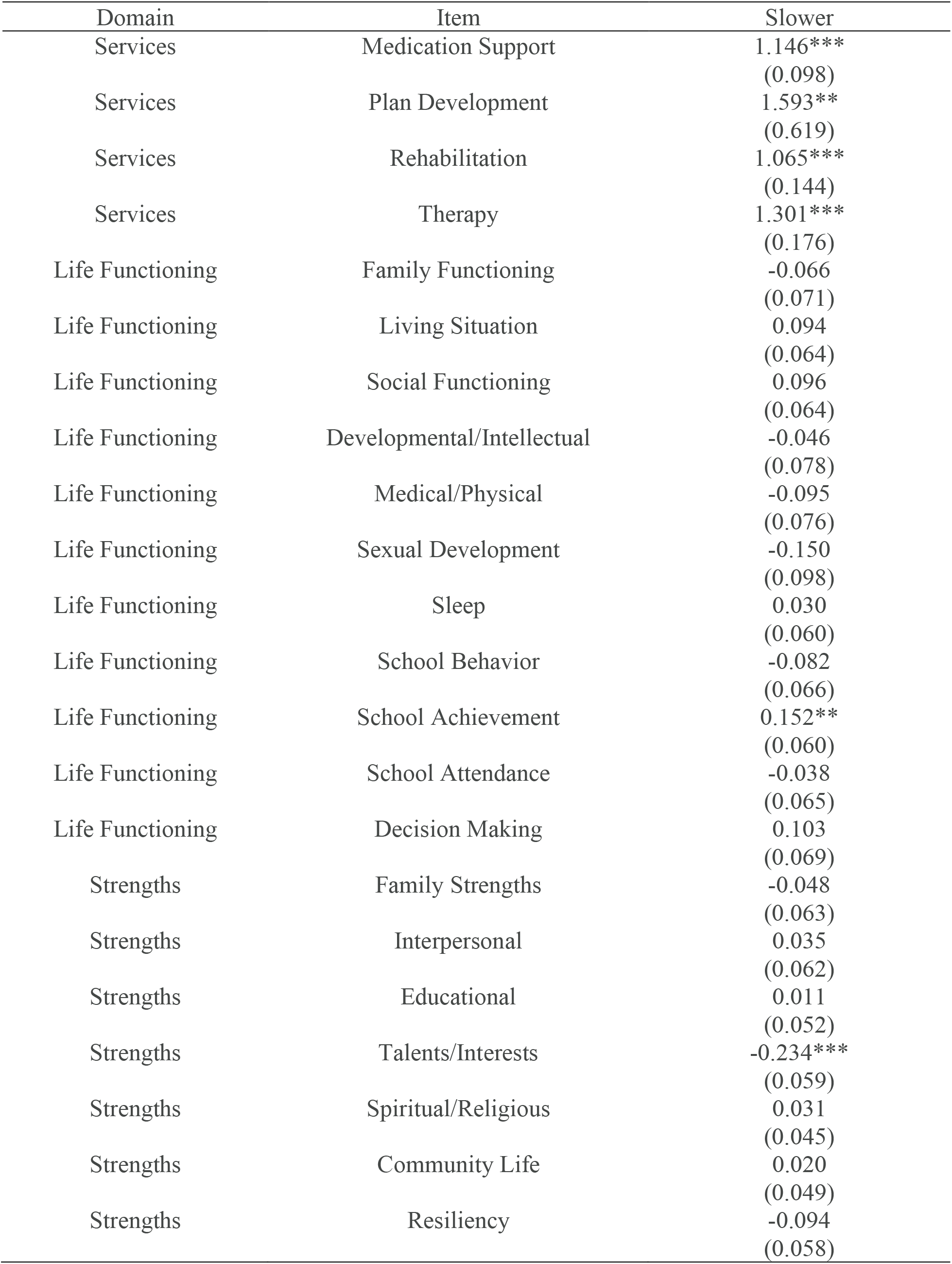
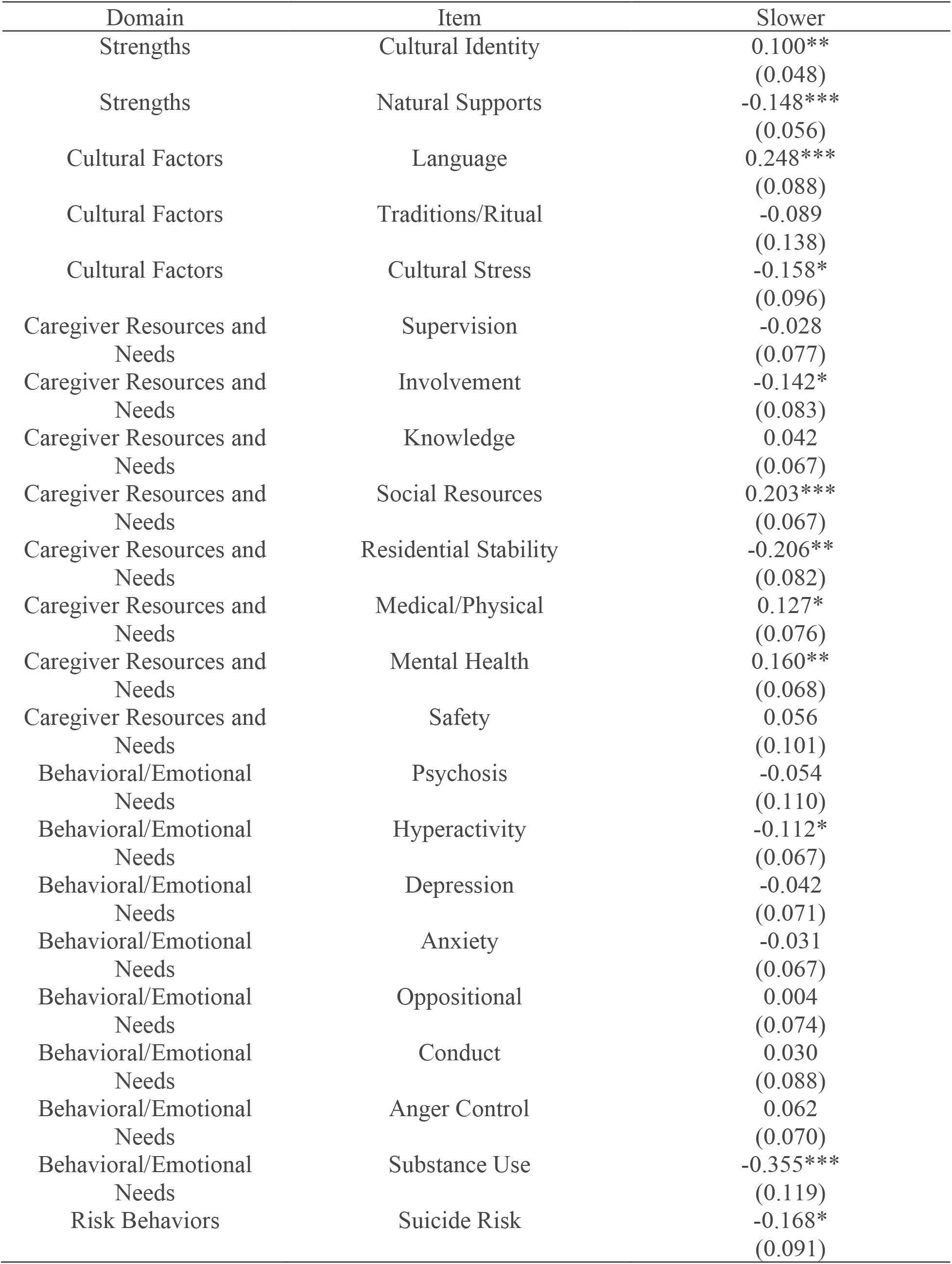
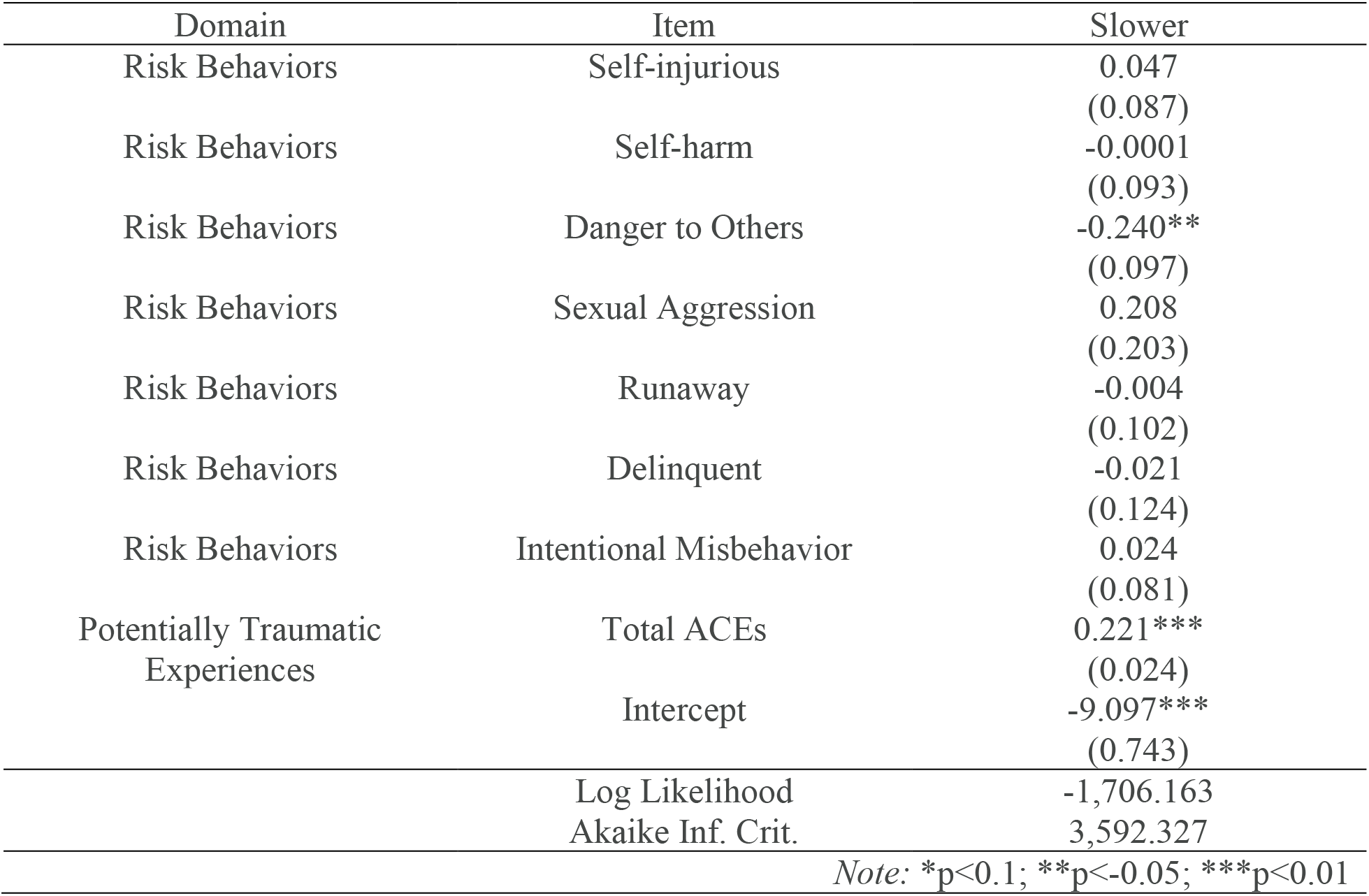

| Domain | Item | Slower |
| --- | --- | --- |
| Potentially Traumatic Experiences | Total ACEs | 0.209***<br>(0.015) |
|  | Intercept | -1.556***<br>(0.064) |
|  | Log Likelihood | -2,362.839 |
|  | Akaike Inf. Crit. | 4,729.677 |
| *Note: \*p<0.1; \*\*p<0.05; \*\*\*p<0.01* |  |  |
| Sociodemographics | Age | -0.069***<br>(0.017) |
| Sociodemographics | Female | -0.015<br>(0.097) |
| Sociodemographics | Black | 0.036<br>(0.165) |
| Sociodemographics | White | 0.199<br>(0.141) |
| Sociodemographics | Hispanic | 0.097<br>(0.114) |
| County | Fresno | 0.386**<br>(0.190) |
| County | Placer | -0.245<br>(0.275) |
| County | Riverside | -11.813<br>(519.846) |
| County | Sacramento | -0.350<br>(0.238) |
| County | San Bernardino | -1.108***<br>(0.235) |
| County | Santa Clara | -0.673***<br>(0.199) |
| County | Solano | -1.335***<br>(0.460) |
| County | Tulare | -11.657<br>(620.609) |
| Diagnosis | Depressive | 1.562***<br>(0.183) |
| Diagnosis | Anxiety | 1.483***<br>(0.200) |

|  | Item | Slower |
| --- | --- | --- |
| Diagnosis | Disruptive Behavior | 1.442***<br>(0.203) |
| Diagnosis | Trauma and Stress | 1.044***<br>(0.180) |
| Diagnosis | Neurodevelopmental | 1.816***<br>(0.218) |
| Diagnosis | Bipolar | 1.156**<br>(0.457) |
| Diagnosis | Psychotic | 1.556***<br>(0.585) |
| Diagnosis | Obsessive Compulsive | 0.820<br>(0.855) |
| Diagnosis | Feeding and Eating | -11.347<br>(594.700) |
| Diagnosis | Personality Disorder | 0.087<br>(1.392) |
| Diagnosis | Substance Related | -10.854<br>(882.743) |
| Programs | Outpatient | 1.195***<br>(0.147) |
| Programs | Prevention | 0.723*<br>(0.370) |
| Programs | Traditional Wraparound | 0.422**<br>(0.190) |
| Programs | Intensive In-home | 0.669***<br>(0.147) |
| Programs | School-based Services | 0.470**<br>(0.221) |
| Programs | Therapeutic Behavioral Services | 0.497<br>(0.340) |
| Programs | 0-5 Programs | 0.498*<br>(0.280) |
| Programs | Crisis Programs | -0.652***<br>(0.210) |
| Programs | Foster Care | -0.310<br>(0.675) |
| Programs | Co-occurring Treatment | 1.319***<br>(0.379) |
| Services | Assessment | 1.712***<br>(0.253) |
| Services | Case Management | 0.792***<br>(0.143) |
| Services | Crisis | 0.718***<br>(0.206) |
| Services | Medication Support | 1.146***<br>(0.098) |
| Services | Plan Development | 1.593**<br>(0.619) |
| Services | Rehabilitation | 1.065***<br>(0.144) |
| Services | Therapy | 1.301***<br>(0.176) |
| Life Functioning | Family Functioning | -0.066<br>(0.071) |
| Life Functioning | Living Situation | 0.094<br>(0.064) |
| Life Functioning | Social Functioning | 0.096<br>(0.064) |
| Life Functioning | Developmental/Intellectual | -0.046<br>(0.078) |
| Life Functioning | Medical/Physical | -0.095<br>(0.076) |
| Life Functioning | Sexual Development | -0.150<br>(0.098) |
| Life Functioning | Sleep | 0.030<br>(0.060) |
| Life Functioning | School Behavior | -0.082<br>(0.066) |
| Life Functioning | School Achievement | 0.152**<br>(0.060) |
| Life Functioning | School Attendance | -0.038<br>(0.065) |
| Life Functioning | Decision Making | 0.103<br>(0.069) |
| Strengths | Family Strengths | -0.048<br>(0.063) |
| Strengths | Interpersonal | 0.035<br>(0.062) |
| Strengths | Educational | 0.011<br>(0.052) |
| Strengths | Talents/Interests | -0.234***<br>(0.059) |
| Strengths | Spiritual/Religious | 0.031<br>(0.045) |
| Strengths | Community Life | 0.020<br>(0.049) |
| Strengths | Resiliency | -0.094<br>(0.058) |
| Strengths | Cultural Identity | 0.100**<br>(0.048) |
| Strengths | Natural Supports | -0.148***<br>(0.056) |
| Cultural Factors | Language | 0.248***<br>(0.088) |
| Cultural Factors | Traditions/Ritual | -0.089<br>(0.138) |
| Cultural Factors | Cultural Stress | -0.158*<br>(0.096) |
| Caregiver Resources and<br>Needs | Supervision | -0.028<br>(0.077) |
| Caregiver Resources and<br>Needs | Involvement | -0.142*<br>(0.083) |
| Caregiver Resources and<br>Needs | Knowledge | 0.042<br>(0.067) |
| Caregiver Resources and<br>Needs | Social Resources | 0.203***<br>(0.067) |
| Caregiver Resources and<br>Needs | Residential Stability | -0.206**<br>(0.082) |
| Caregiver Resources and<br>Needs | Medical/Physical | 0.127*<br>(0.076) |
| Caregiver Resources and<br>Needs | Mental Health | 0.160**<br>(0.068) |
| Caregiver Resources and<br>Needs | Safety | 0.056<br>(0.101) |
| Behavioral/Emotional<br>Needs | Psychosis | -0.054<br>(0.110) |
| Behavioral/Emotional<br>Needs | Hyperactivity | -0.112*<br>(0.067) |
| Behavioral/Emotional<br>Needs | Depression | -0.042<br>(0.071) |
| Behavioral/Emotional<br>Needs | Anxiety | -0.031<br>(0.067) |
| Behavioral/Emotional<br>Needs | Oppositional | 0.004<br>(0.074) |
| Behavioral/Emotional<br>Needs | Conduct | 0.030<br>(0.088) |
| Behavioral/Emotional<br>Needs | Anger Control | 0.062<br>(0.070) |
| Behavioral/Emotional<br>Needs | Substance Use | -0.355***<br>(0.119) |
| Risk Behaviors | Suicide Risk | -0.168*<br>(0.091) |
| Risk Behaviors | Self-injurious | 0.047<br>(0.087) |
| Risk Behaviors | Self-harm | -0.0001<br>(0.093) |
| Risk Behaviors | Danger to Others | -0.240**<br>(0.097) |
| Risk Behaviors | Sexual Aggression | 0.208<br>(0.203) |
| Risk Behaviors | Runaway | -0.004<br>(0.102) |
| Risk Behaviors | Delinquent | -0.021<br>(0.124) |
| Risk Behaviors | Intentional Misbehavior | 0.024<br>(0.081) |
| Potentially Traumatic<br>Experiences | Total ACEs | 0.221***<br>(0.024) |
|  | Intercept | -9.097***<br>(0.743) |
|  | Log Likelihood | -1,706.163 |
|  | Akaike Inf. Crit. | 3,592.327 |
| *Note: \*p<0.1; \*\*p<0.05; \*\*\*p<0.01* |  |  |

## Appendix 4. Strengths over time

|  | High Initial | Low Initial | High Final | Low Final |
| --- | --- | --- | --- | --- |
| Faster | 51.69% | 53.25% | 55.74% | 50.08% |
| Slower | 19.30% | 23.50% | 23.42% | 20.84% |
| Minimal | 29.01% | 23.25% | 20.84% | 29.08% |

